# An RNA biomarker panel for the diagnosis of Alzheimer’s disease from whole blood

**DOI:** 10.1101/2025.08.04.25332553

**Authors:** Sean Paz, Janet D. Robishaw, Massimo Caputi

## Abstract

Alzheimer’s Disease (AD) diagnosis remains primarily clinical, relying on patient history, cognitive tests, and sometimes brain imaging. Emerging approaches include advanced neuroimaging techniques and the use of protein biomarkers in cerebrospinal fluid and plasma. However, these methods have limitations such as high costs and the need for specialized infrastructures. In contrast, analysis of RNA expression in blood samples could be achieved with a high degree of sensitivity and accuracy in point of care settings, providing an ideal platform for a timely and accurate AD diagnosis. Peripheral blood samples from 50 AD patients and 50 healthy subjects were analyzed utilizing a whole transcriptome RNA sequencing approach to determine differences in the expression of i) mRNAs, ii) lncRNAs, iii) miRNAs, iv) circRNAs and v) alternatively spliced mRNA isoforms. Multiple parallel analysis pipelines were utilized to identify four differentially expressed transcripts to form an AD biomarker panel. Expression levels in each AD patient were compared to those in healthy subjects to obtain an AD predictive score (ADps) with a positive predictive value of over 90% and a specificity of over 95%. The four RNA biomarkers can reliably identify AD patients using a minimal amount of peripheral blood and has the potential to be developed into a minimally invasive, cost-effective and reliable tool for early AD diagnosis.

## Introduction

Dementia is a clinical syndrome caused by various conditions affecting the brain and is characterized by cognitive decline that interferes with daily functioning. As of 2023, an estimated 55 million people worldwide are living with dementia, with this number projected to rise to 78 million by 2030 [1]. In the United States, Alzheimer’s disease (AD) is the most common form of dementia, affecting approximately 7 million Americans aged 65 and older; this number is expected to increase to 14 million by 2060 [2]. The increasing prevalence of dementia, and specifically AD, underscores the urgent need for research and healthcare planning to address its growing impact on individuals and society.

Early detection and diagnostic accuracy are essential for enabling timely intervention and treatment of AD. However, current diagnostic strategies are largely clinical and typically occur only after noticeable symptoms and cognitive decline have already manifested. These approaches rely heavily on patient history, cognitive assessments, and, in some cases, conventional brain imaging—methods that often result in delayed diagnosis. To address this gap, newer techniques are being developed to identify AD at much earlier stages, ideally before significant neurodegeneration occurs. These include advanced neuroimaging modalities such as amyloid and tau positron emission tomography (PET) scans, which allow for the in vivo visualization of amyloid-beta plaques and tau tangles in the brain [3]. Additionally, protein biomarkers in cerebrospinal fluid (CSF) and blood—such as phosphorylated tau (p-Tau 181, p-Tau 217, p-Tau 231, MTBR-tau243) and amyloid-beta species ratios (Aβ42/40) are emerging as promising diagnostic indicators when measured using mass spectrometry-based (MS) and fully automated immunoassay platforms [4–8]. Machine learning algorithms further enhance diagnostic potential by integrating multi-modal data, including genetic, imaging, and clinical information, to improve predictive models for AD [9]. Although collectively these techniques show promise to enhance early diagnosis and allow for more effective intervention strategies, they are still mainly used in research settings and have several drawbacks. PET scans are highly specific, but their utility is limited by high costs, requirement for highly specialized infrastructure and only provide information on amyloid or tau-dependent pathologies, while the pathophysiology of AD can be multifactorial and highly complex. Conversely, CSF-based biomarkers present a cost-effective approach to the evaluation of multiple markers. However, the invasive nature of the lumbar puncture limits their application. Blood based biomarkers have the potential to provide an affordable diagnosis at the point of care, although the quantitative measurement of specific protein isoforms or modifications, often present in trace amounts, can be reliable when carried out in research laboratories but is often technically challenging in a clinical setting.

Given these limitations, there is growing interest in exploring molecular signatures that could offer earlier, less invasive, and more comprehensive insights. In this context, RNA-based biomarkers have emerged as promising candidates. Recent advances in genomics and RNA sequencing techniques have enabled the study of different classes of RNAs as potential AD biomarkers. Coding (mRNAs) and non-coding RNAs, such as microRNA (miRNAs) and circular RNAs (circRNAs), have emerged as promising biomarkers thanks to their roles in the regulation of gene expression and cellular processes implicated in neurodegeneration [10, 11]. Mature miRNAs regulate gene expression by binding to the 3’-untranslated region (3’UTR) of mRNAs and blocking protein synthesis or leading to the degradation of the target mRNAs [12]. miRNAs have been shown to be dysregulated in AD patients, influencing processes related to amyloid-beta processing, tau phosphorylation, and inflammation [13, 14]. circRNAs are less susceptible to degradation than canonical linear RNAs [15] and often act as sponges for miRNAs, thus inhibiting their functions, and can be differentially expressed in AD patients [16, 17]. Similarly, the expression of long non-coding RNAs (lncRNAs), like BACE1-AS, which regulates the expression of the β-site amyloid precursor protein cleaving enzyme 1 (BACE1), is altered in AD and correlates with disease progression [18]. Overall, RNA molecules could provide insights into the molecular mechanisms underlying AD and offer potential for the development of diagnostic tools and therapeutic strategies.

Unlike previous studies that have focused on specific RNA classes, mostly miRNAs or mRNAs, in cell free CNS or plasma samples, our study takes a broader approach by analyzing all classes of RNAs present in whole blood. To date, research utilizing whole blood has been extremely limited [19–22]. In this study we analyzed blood samples from 50 AD patients and 50 healthy individuals of similar age, sex and ethnic background utilizing a whole transcriptome RNA sequencing (RNASeq) approach to determine differences in the expression of several classes of coding and non-coding RNAs.

## Methods

### Patients

Whole blood samples from AD patients (n=50) and healthy control subjects (n=50) were obtained from a commercial biobank (PrecisionMed). Samples were de-identified and are exempt from Institutional Review Board (IRB) approval. AD patients were defined by a clinical diagnosis carried out by neurologists in accordance with criteria adopted by the National Institute of Neurological and Communicative Disorders and Stroke Alzheimer’s Disease and Related Disorders Association (NINCDS-ADRDA) supported by a Mini-Mental State Examination (MMSE) score <25, and an Alzheimer’s Disease Assessment Scale - Cognitive Subscale (ADAS-Cog) score >13 and did not have other remarkable clinical history. Healthy donors exhibited an unremarkable clinical history, and their cognitive test scores were within normal ranges (MMSE >28). Variable ages, both sexes and heterogeneous ethnic backgrounds were present in both AD and control patients (summarized in Fig. 1A and Supplemental Data Table S1). Peripheral whole blood samples were drawn by venipuncture and preserved in PAXgene RNA tubes (Bekton Dickinson) at -80C, according to manufacturer instructions for long term storage and shipping.

**Figure 1.**
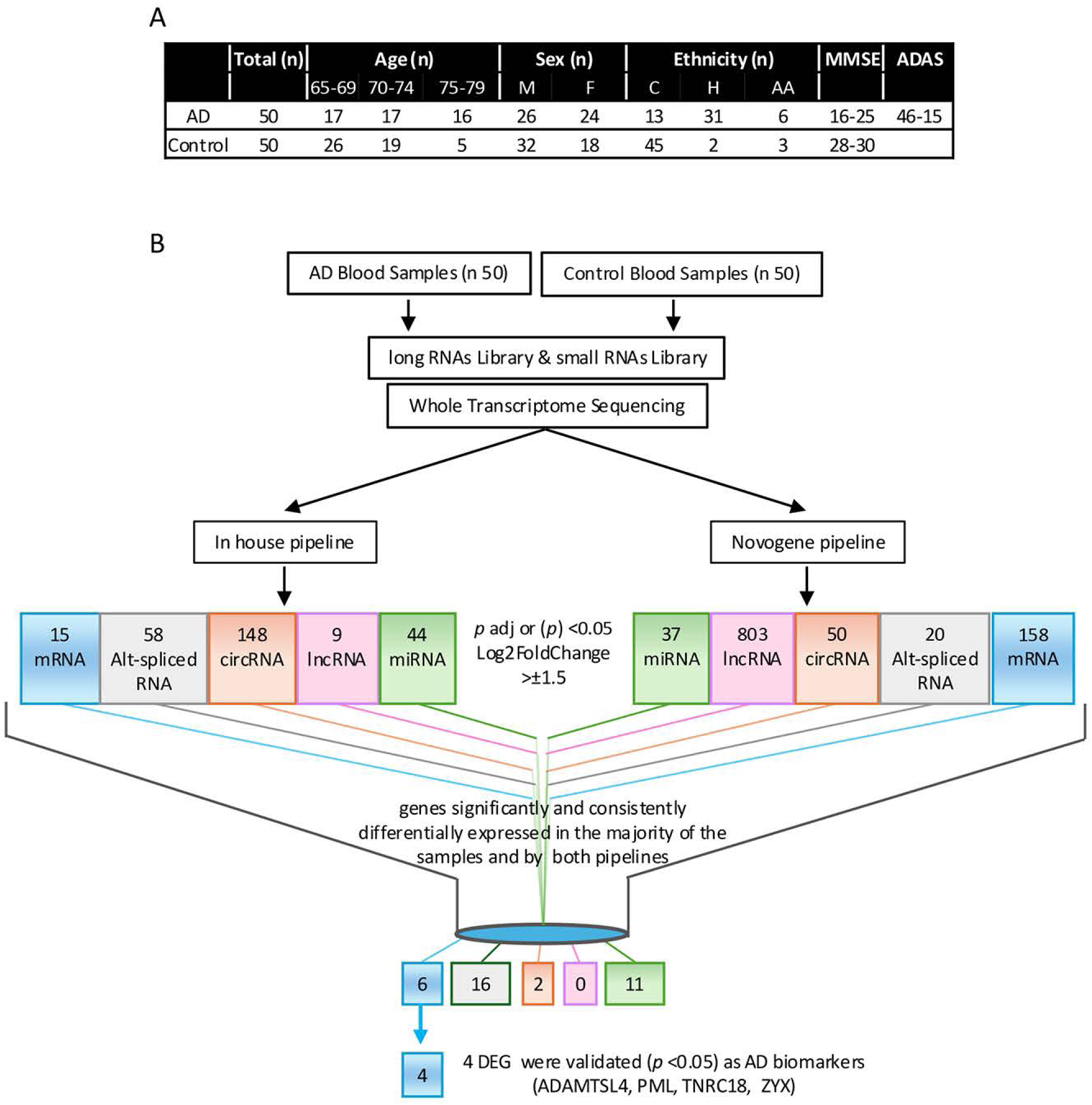
Whole transcriptome RNA sequencing. (A) Clinical characteristics of AD patients and control subjects. Age group, sex and ethnic background (C, Caucasian; H, Hispanic; AA, African American) are reported for AD and control patients. Score intervals for Mini Mental State Examination (MMSE) and Alzheimer’s Disease Assessment Scale - Cognitive Subscale (ADAS-Cog) are also reported. (B) Workflow of the whole transcriptome RNA sequencing analysis of peripheral blood samples from AD patients and healthy control subjects. Total RNA extracted from AD patients and controls was utilized to generate long and short RNA libraries. Reads generated from the sequencing of both libraries were analyzed using 2 independent pipelines to determine and prioritize DE miRNAs, lncRNAs, circRNAs, mRNAs and alternatively spliced RNAs. 35 transcripts were selected for the validation step, which yielded the final 4 RNA biomarker panel.

### RNA isolation, cDNA libraries and transcriptome sequencing

Total RNA was extracted utilizing a TRIzol (Thermo-Fisher Scientific) based protocol optimized for the extraction of limiting RNA amounts [23], treated with Turbo DNase (Thermo-Fisher Scientific). PAXgene tubes were thawed at room temperature on a rotator for two hours. Samples were then centrifuged for 10 minutes at 3,000 x g, the supernatant was discarded and 4mL of ddH2O was added to the cell pellet resuspend by vortexing. Samples were centrifuged for 10 minutes at 3,000 x g, the supernatant was discarded and 1mL of TRIzol was added and the pellet was resuspended by vortexing. Samples were incubated at room temperature for 10 minutes, added to a QIAshredder column (Qiagen) and centrifuged for 2 minutes at 16,000 x g. The eluate was transferred to a new microcentrifuge tube and 200µL of chloroform was added. Samples were vortexed, allowed to separate for 3 minutes and centrifuged at 16,000 x g for 15 minutes at 4°C. The top aqueous phase was transferred into a fresh microcentrifuge tube, 50µg of glycogen and 1mL of 100% ethanol were added. Samples were incubated at -80°C for 30 minutes and then centrifuged at 16,000 x g for 25 minutes at 4°C. The supernatant was decanted, and the pellet was washed with 75% ethanol and centrifuged at 16,000 x g for 5 minutes at 4°C. The supernatant was aspirated, the pellet air dried and then resuspended in 40µL of ddH2O. RNA quality was analyzed utilizing an Agilent Tapestation 4200 and RNA integrity (RIN) was maintained at ≥7. RNA concentration and purity were measured using a NanoDrop 2000 (Thermo Scientific). The total RNA extracted from each sample was partitioned into 3 aliquots. One aliquot was utilized for the preparation of the Illumina TruSeq RNA Library Prep Kit v2 (Illumina) following rRNA removal using the Globin-Zero Gold rRNA Removal Kit (illumina). The second aliquot of RNA was utilized for the preparation of the TruSeq Small RNA Library Prep Kit (Illumina). The third aliquot was utilized for the RT-PCR and RT-qPCR validation assays. Pooled samples used in the first validation step were obtained by combining 100 ng of RNA extracted from 8 groups of 5 AD or 5 control donors as indicated in Additional Data 1 (Table S1). The libraries were sequenced utilizing an NovaSeq6000 (Illumina) generating 40 million 150 nt paired-end reads (long RNA) and 20 million 50 nt single-end reads (short RNA). Library preparations and sequencing were performed by Novogene Ltd.

### Raw sequence reads analysis

The raw read pre-processing and downstream analysis were carried out utilizing 2 parallel pipelines that utilize different sets of tools to determine the differential expression of: i) protein coding mRNAs, ii) long non-coding RNAs (lncRNAs), iii) miRNAs, iv) circRNAs and v) alternatively spliced RNA isoforms (summarized in Fig. S2). One pipeline was set-up by our in-house bio-computing group at Florida Atlantic University, while a second pipeline was provided by Novogene. Raw reads underwent quality control, including adaptor trimming and low-quality read filtering, prior to alignment to GRCh38 human genome assembly. All sequencing data are publicly available on the GEO database (GSE248423). Transcripts with an adjusted *p*-value <0.05 and ∣log2foldchange(FC)| >1.5 or ∣InclusionLevelDifference| >0.15 were prioritized as differentially expressed (DE) mRNAs, DE lncRNAs, DE miRNAs, DE circRNAs and alternatively spliced mRNAs (shown in source data table). Targets prioritized by both pipelines were curated to eliminate transcripts with low expression levels (TPM or FPKM >1) or displaying expression changes only in a minority of the samples and selected for validation.

### RT-PCR and RT-qPCR

RNA was reverse transcribed utilizing a random pd(N)6 primer and Superscript II RT (Thermo-Fischer Scientific). PCR assays for the amplification of the alternatively spliced isoforms were carried out utilizing the 3G HotStart Taq DNA Polymerase (BioBasic) for 35 cycles (95°C 30 s, 60°C 1 min, 72°C 1 min) and a slow renaturation step added after the last cycle to minimize the formation of hybrid PCR products between alternatively spliced isoforms as previously described [24]. PCR products were separated on a 1.5 % agarose gel and quantified utilizing an Odyssey classic imaging system (LICOR Biosciences). Quantitative PCR (qPCR) assays for the amplification of mRNA, lncRNA and circRNAs were performed utilizing, the Green-2-Go SYBR green qPCR Kit (BioBasic), according to the manufacturer instructions. qPCR assays were carried out utilizing the reaction conditions: 95°C 10 min, followed by 40 cycles: 95°C 15s and 62°C 60s utilizing an Agilent AriaMx real time PCR system. Cycle thresholds (Cts) values were normalized for the relative expression of the housekeeping gene RPL13A and analyzed with the AriaMx software. Quantitative PCR analysis of miRNAs was carried out utilizing the Mir-X™ miRNA First-Strand Synthesis and miRNA qRT-PCR TB Green Kits (Takara Bio). Cts values were normalized for the expression of the small RNA (snRNA) U6. All qPCR assays were run with technical duplicates. Primer sequences are listed in Supplemental Data Table S2. Primers amplifying the circRNA substrates are designed to specifically amplify circRNAs but not linear RNAs since they extend in opposite directions on linear transcripts but extend towards each other on the circularized RNAs. The qPCR denaturation profiles for all the primer pairs utilized in this study showed single, distinct denaturation peaks. Average Ct values of the technical duplicates before and after normalization for RPL13 or U6 snRNA expression, are shown in the supplemental source data.

### Statistical analysis

Differences in transcript expression between control and AD patients were compared using Welch’s t-test (first validation step, pooled RNA samples). The Mann-Whitney U test was used when the distribution was skewed (Fig. 3A, S2). In the box and whisker plot: error bars represent the 95% confidence interval, the bottom and top of the box are the 25th and 75th percentiles, respectively, the line inside the box is the 50th percentile (median), and outliers are shown as circles.

**Figure 2.**
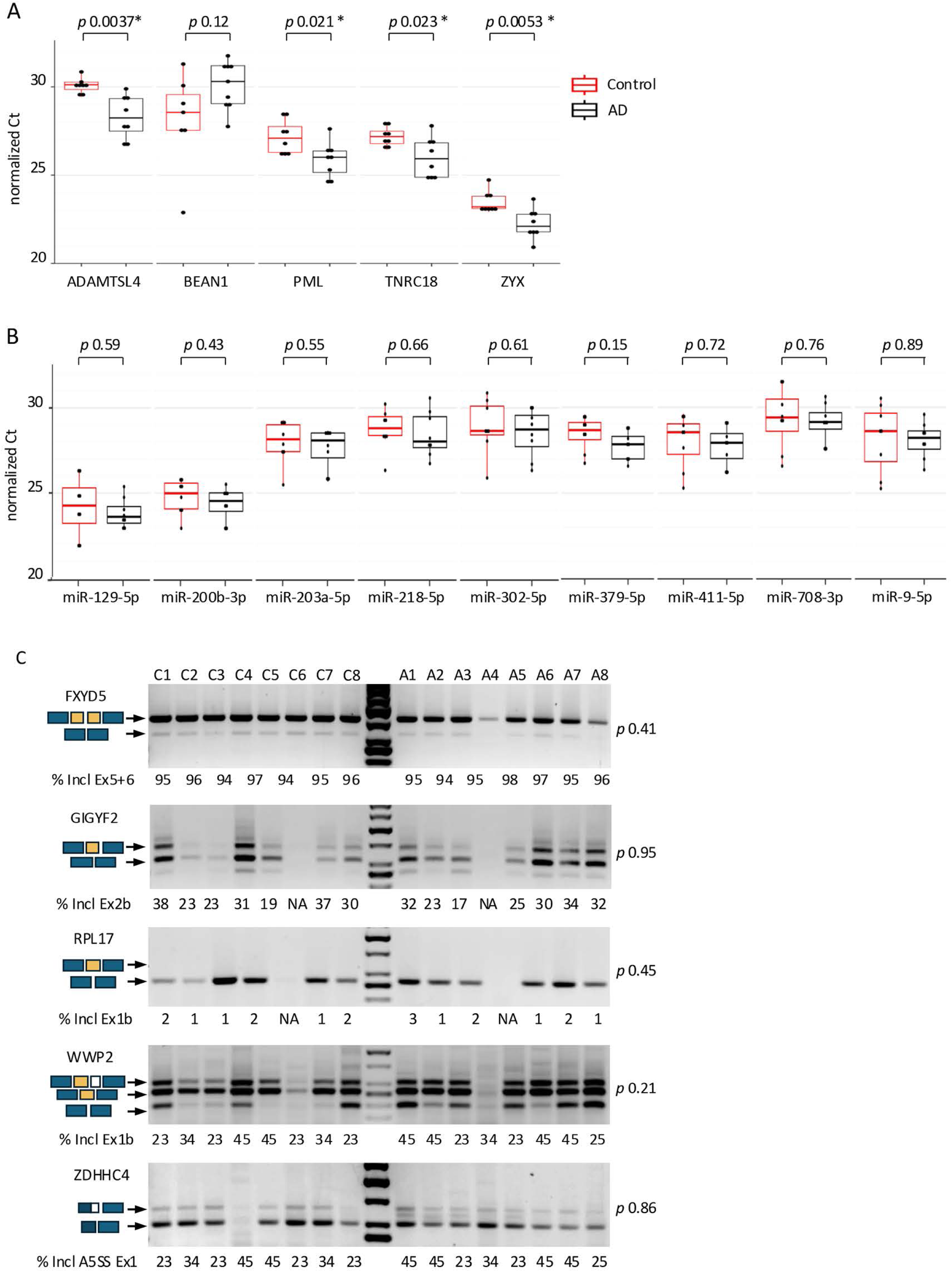
Validation of transcripts prioritized by the data analysis. (A) Boxplots showing normalized Ct values for 6 mRNAs and (B) 9 miRNAs selected for validation and efficiently amplified by RT-qPCR from the total RNA extracted from the peripheral blood of 40 AD patients and 40 healthy subjects RNA samples were pooled into 8 AD and 8 control samples. (C) RT-PCR analysis of the alternative splicing patterns of the 5 selected transcripts that amplify efficiently RNA from the pooled samples. Alternatively spliced exons are indicated on the left, and the percentage of exon inclusion is indicated underneath each sample. Control samples (Cn), AD samples (An). \**p* value (Welch’s t-test) <0.05 was considered significant.

**Figure 3.**
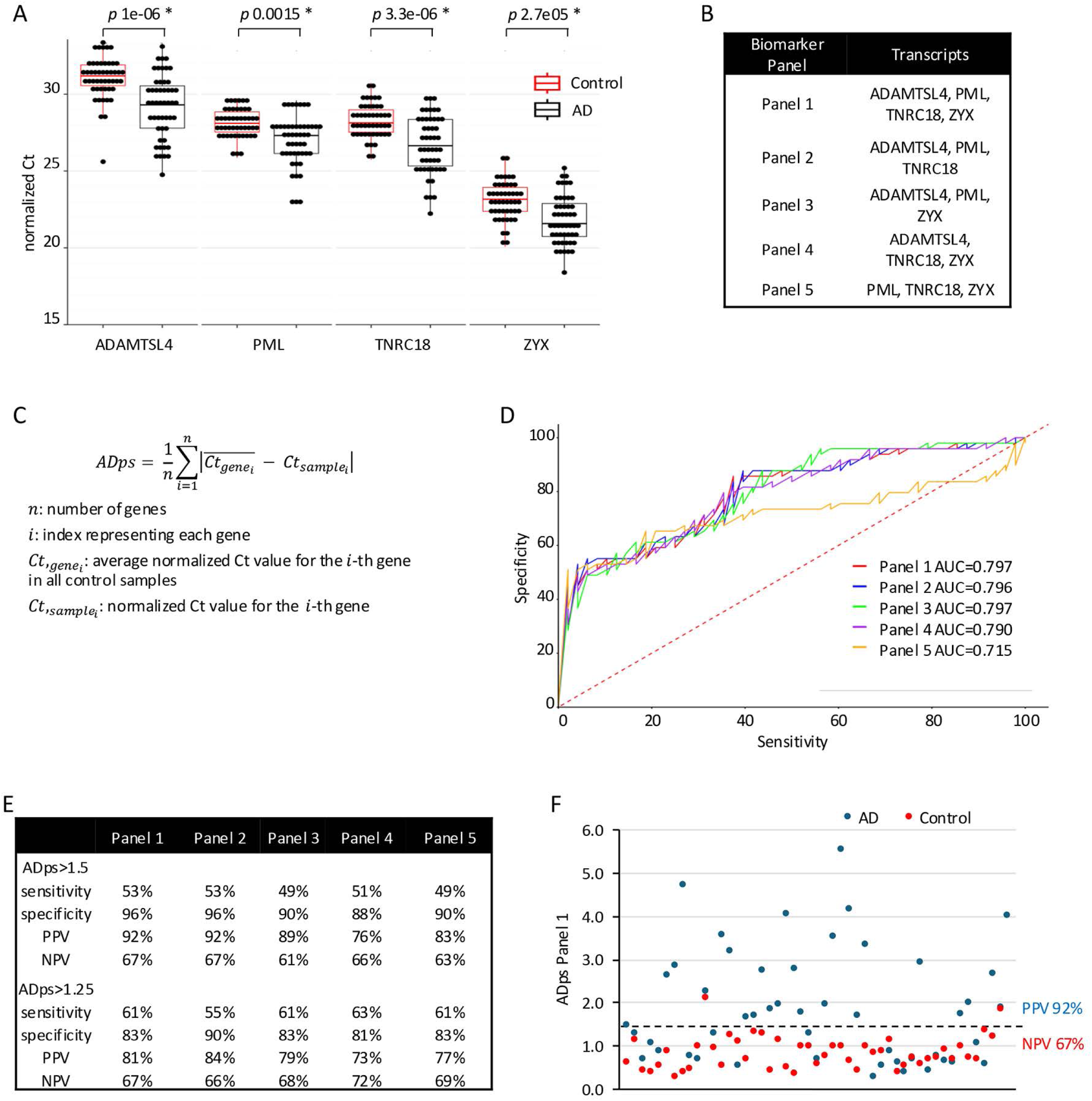
A 4 biomarker RNA panel. (A) Boxplots of the normalized Ct values for the 4 biomarker RNAs: ADAMTSL4, PML, TNRC18, and ZYX, obtained by amplifying the total RNA extracted from 50 AD patients and 50 healthy control subjects by RT-qPCR. * *p* value (Mann-Whitney U test) <0.05 was considered significant. (B) The AD predictive score (ADps) is calculated combining the normalized Ct values of each RNA biomarker in each sample analyzed and comparing them with their normalized Ct values in a group of control samples. Receiver operating characteristic (ROC) and the area under the curve (AUC) was used to assess the sensitivity and specificity of the different combination of transcripts used to calculate the ADps score. (C) Diagnostic performance parameters including sensitivity, specificity, positive and negative predictive values (PPV, NPV) for two ADps thresholds (>1.5 and >1.25) and 5 different combinations of RNAs biomarkers. (D) ADPs for each AD and control sample utilizing the 4 RNA biomarkers AD panel. PPV and NPV for an ADps threshold >1.5 are shown.

AD predictive scores (ADps) were calculated as the average absolute deviation between the average normalized Ct value of each biomarker RNA in all control samples and their normalized Ct value in each sample (Fig. 3B). Receiver operating characteristic (ROC) and the area under the curve (AUC) was used to test for sensitivity and specificity of different combination of transcripts utilized to generate the ADps score. Spearman correlation analyses were used to determine the correlation between ADps score and age or MMSE score. 2-way ANOVA was utilized to determine correlations between ADps race and condition while Mann-Whitney U test was utilized to determine correlation between ADps sex and condition. A p-value <0.05 was considered statistically significant. All statistical analyses in this study were performed using R (version 4.4.0; https://www.r-project.org/).

### Code availability

The following sets of processing tools and suites were utilized in this study HISAT2[25], Cufflinks [26], rMATs [27], miRDeep2 [28], Cutadapt [29], FastQC [30], CIRCexplorer2 [31], CIRIquant [32], DESeq2 [33], EdgeR [34].

## Results

### Whole transcriptome analysis

We utilized a whole transcriptome RNA sequencing (RNASeq) approach to investigate the transcriptome of peripheral blood samples from AD patients (n=50) and healthy donors (n=50) (Figure 1). AD patients were defined by a clinical diagnosis supported by a Mini-Mental State Examination (MMSE) score of <25, and an Alzheimer’s Disease Assessment Scale - Cognitive Subscale (ADAS-Cog) score of >13. Healthy control subjects exhibited an unremarkable clinical history, and their cognitive tests scores were within normal ranges (MMSE >28). Both AD and control groups included individuals of varying ages, both sexes, and heterogeneous ethnic backgrounds (Fig. 1A and Tab. S1).

Total RNA extracted from each sample was partitioned into 3 aliquots. Long and short cDNA libraries were synthesized using two of the aliquots, while the third aliquot was utilized in validation assays. RNASeq assays carried out utilizing the long cDNA library generated 40 million 150 nt paired*-*end reads while the short cDNA library generated 20 million 50 nt single-end reads. The RNAseq datasets were analyzed utilizing two independent analysis pipelines (in-house and Novogene, Fig. S1) to determine with a high degree of confidence the differential expression of: i) protein coding mRNAs, ii) long non-coding RNAs (lncRNAs), iii) miRNAs, iv) circRNAs and v) alternatively spliced mRNA isoforms (Alt Spl) (Fig 1B).

Differentially expressed (DE) transcripts in all analysis pipelines were prioritized utilizing a significant false discovery rate (FDR) adjusted *p*-value <0.05 and differences in expression changes (∣Log2FoldChange∣) >1.5 or alternative splicing differences (∣InclusionLevelDifference∣) >0.15. Targets common to the prioritized lists from both, in-house and Novogene, pipelines were further filtered to eliminate DE transcripts with low expression levels (TPM or FPKM > 1) or displaying expression changes in only a minority of samples. This yielded a curated set of 35 DE and Alt Sp1 transcripts, including 6 coding mRNAs, 11 miRNAs, 2 circRNAs, and 16 alternatively spliced isoforms, which were then selected for downstream experimental validation (Table 1).

**Table 1.** RNAs selected for validation. Gene names corresponding to each RNA are listed along with log2 fold changes and adjusted *p* values from both pipelines for DE RNAs. For alternatively splice RNAs, the Inclusion Level Difference (ILD) is reported. ‡ Type of change observed: differentially expressed (DE), skipped exon (SE), alternative 5’ splice site (A5SS), alternative 3’ splice site (A3SS), mutually exclusive exons (MXE), and retained intron (RI) and transcript exon number are indicated. * mir302a-3p, mir302b-3p, mir302c-3p, mir302d-3p differ from each other only by a single nucleotide and can be detected by a common primer set. Transcripts showing a robust amplification by either RT-PCR or RT-qPCR from RNA extracted from blood samples are indicated.

| DE RNA | description | RNA type | change ‡ | log2_FC InHouse | log2_FC Novogene | Adj_p InHouse | Adj_P Novogene | validation assay | detected in blood |
| --- | --- | --- | --- | --- | --- | --- | --- | --- | --- |
| ADAMTSL4 | ADAMTS like 4 | mRNA | DE | 2.05 | 1.56 | 1.80E-03 | 2.40E-16 | RT-qPCR | YES |
| BEAN1 | brain expressed associated with NEDD4 1 | mRNA | DE | 1.7 | 1.92 | 1.80E-03 | 2.10E-03 | RT-qPCR | YES |
| PEAK3 | PEAK family member 3 | mRNA | DE | 1.86 | 1.64 | 2.10E-02 | 2.10E-08 | RT-qPCR | NO |
| PML | PML nuclear body scaffold | mRNA | DE | 1.57 | 1.17 | 1.80E-03 | 1.90E-09 | RT-qPCR | YES |
| TNRC18 | trinucleotide repeat containing 18 | mRNA | DE | 1.5 | 1.36 | 1.80E-03 | 1.80E-17 | RT-qPCR | YES |
| ZYX | zyxin | mRNA | DE | 1.52 | 1.45 | 1.80E-03 | 1.30E-15 | RT-qPCR | YES |
| miR-9-5p |  | miRNA | DE | -2.52 | -2.5 | 1.20E-06 | 2.40E-06 | RT-qPCR | YES |
| miR-129-5p |  | miRNA | DE | -1.57 | -1.67 | 1.50E-06 | 1.10E-06 | RT-qPCR | YES |
| miR-200b-3p |  | miRNA | DE | -3.32 | -3.91 | 1.00E-19 | 9.50E-30 | RT-qPCR | YES |
| miR-203a-3p |  | miRNA | DE | -1.76 | -1.89 | 3.40E-05 | 2.40E-05 | RT-qPCR | YES |
| miR-218-5p |  | miRNA | DE | -1.92 | -1.99 | 4.50E-04 | 5.70E-04 | RT-qPCR | YES |
| miR-302a-3p* |  | miRNA | DE | -5.14 | -5.16 | 8.20E-18 | 5.60E-18 | RT-qPCR | YES |
| miR-302b-3p* |  | miRNA | DE | -5.7 | -5.84 | 2.30E-23 | 5.50E-23 | RT-qPCR | YES |
| miR-302d-3p* |  | miRNA | DE | -4.9 | -4.85 | 1.20E-13 | 3.50E-13 | RT-qPCR | YES |
| miR-379-5p |  | miRNA | DE | -2.33 | -2.32 | 1.70E-10 | 1.90E-09 | RT-qPCR | YES |
| miR-411-5p |  | miRNA | DE | -2.4 | -2.3 | 1.30E-06 | 2.40E-06 | RT-qPCR | YES |
| miR-708-3p |  | miRNA | DE | -4.13 | -4.18 | 3.50E-14 | 2.20E-14 | RT-qPCR | YES |
| FAM135A_0007 |  | circRNA | DE | -1.7 | -1.86 | 3.60E-03 | 8.80E-03 | RT-qPCR | NO |
| SPATA13_0008 |  | circRNA | DE | -151 | -180 | 7.98E-03* | 2.77E-02* | RT-qPCR | NO |
| Alt Spl RNA | description | gene type | change ‡ | ILD InHouse | ILD Novogene | FDR InHouse | p Novogene | validation assay | detected in blood |
| ATG16L2 | autophagy related 16 Like 2 | mRNA | A3SS Ex5 | -0.16 | -0.16 | 9.70E-08 | 4.30E-06 | RT-PCR | NO |
| CFAP44 | cilia and flagella associated protein 44 | mRNA | MXE Ex24+22 | 0.16 | 0.15 | 3.47E-02 | 1.60E-02 | RT-PCR | NO |
| FXYD5 | FXYD domain containing ion transport regulator 5 | mRNA | MXE Ex5+6 | 0.16 | 0.16 | 2.00E-03 | 3.00E-04 | RT-PCR | YES |
| GIGYF2 | GRB10 interacting GYF protein 2 | mRNA | SE Ex2b | -0.18 | 0.18 | 1.15E-02 | 1.90E-02 | RT-PCR | YES |
| KDM2B | lysine demethylase 2B | mRNA | MXE<br>Ex7+9 | 0.21 | 0.21 | 2.77E-05 | 1.20E-05 | RT-PCR | NO |
| MBD6 | methyl-CpG binding domain<br>protein 6 | mRNA | RI In2 | 0.16 | 0.18 | 2.05E-02 | 4.60E-02 | RT-PCR | NO |
| PICALM | phosphatidylinositol binding<br>clathrin assembly protein | mRNA | SE<br>Ex19b | 0.22 | 0.22 | 4.71E-04 | 2.90E-05 | RT-PCR | NO |
| PIGO | phosphatidylinositol glycan<br>anchor biosynthesis class O | mRNA | SE Ex7 | -0.2 | -0.2 | 1.70E-06 | 9.10E-07 | RT-PCR | NO |
| PILRB | paired immunoglobulin like<br>type 2 receptor beta | mRNA | A3SS<br>Ex5 | 0.17 | 0.18 | 6.68E-04 | 3.60E-05 | RT-PCR | NO |
| RBIS | ribosomal biogenesis factor | mRNA | A5SS<br>Ex2 | -0.18 | -0.19 | 2.76E-04 | 1.70E-05 | RT-PCR | NO |
| RPL17 | ribosomal protein L17 | mRNA | SE<br>Ex1b | -0.21 | -0.22 | 2.51E-05 | 1.50E-06 | RT-PCR | YES |
| SNRNP70 | small nuclear<br>ribonucleoprotein U1<br>subunit 70 | mRNA | SE<br>Ex7b | -0.21 | -0.21 | 2.48E-06 | 4.60E-06 | RT-PCR | NO |
| TECPR1 | tectonin beta-propeller<br>repeat containing 1 | mRNA | MXE<br>Ex2+5 | 0.16 | 0.16 | 1.10E-03 | 4.80E-04 | RT-PCR | NO |
| TMEM104 | transmembrane protein 104 | mRNA | MXE<br>Ex3+4 | 0.18 | 0.18 | 1.05E-02 | 4.50E-04 | RT-PCR | NO |
| WWP2 | WW domain containing E3<br>ubiquitin protein ligase 2 | mRNA | SE<br>Ex1b | -0.16 | -0.16 | 1.68E-03 | 2.70E-02 | RT-PCR | YES |
| ZDHHC4 | zinc finger DHHC-type<br>palmitoyltransferase 4 | mRNA | A5SS<br>Ex1 | -0.18 | -0.19 | 9.70E-08 | 1.00E-04 | RT-PCR | YES |

### Validation of the DE RNAs

The DE transcripts prioritized by the parallel data analysis pipelines were validated by RT-qPCR (for DE miRNAs, mRNAs and circRNAs) and RT-PCR (for Alt Sp1 RNAs). Pooled RNA samples from AD and healthy controls were utilized in the initial validation step to reduce the number of single amplification reactions carried out while maintaining the heterogenous range of gene expression observed in individuals. This enhances the robustness of the assay, as reliable disease biomarkers should show consistent differential expression across most of the affected population. 40 AD and 40 control RNA samples were divided into groups of 5 and pooled into 8 AD and 8 control pooled samples.

4 of the 6 differentially expressed coding mRNAs analyzed were significantly differentially expressed (*p* <0.05) in the AD pooled samples when compared to the controls (Fig 2A). One mRNA did not amplify (Ct >35) and was discarded from our analysis since we wanted to identify targets that could be reliably amplified. None of the lncRNAs were selected for validation following our stringent data analysis. And, both circRNAs tested did not amplify within an acceptable Ct range and were also excluded. All selected miRNAs amplified with average Ct values <30 in both AD and control pooled samples; however, none showed significant differential expression between AD and control samples (Fig. 2B). This result was confirmed using individual (non-pooled) samples from 50 AD and 50 control patients for mir-218-5p and mir-379-5p, confirming the robustness of the pooled sample approach (Fig. S1). Of the 13 Alt Sp1 transcripts, only 5 were detectable after a single round of RT-PCR (35 cycles) and none showed significant changes in splicing pattern between the AD and control samples (Fig. 2C). Although it is plausible that some transcripts could be amplified upon further optimization, such as increased PCR cycles or nested PCR, these approaches might compromise data reliability and are not aligned with our criteria for robust AD biomarker identification. Ultimately, 4 coding mRNAs, ADAMTSL4, PML, ZYX and TNRC18, were selected for inclusion in the AD biomarker panel, since they amplified consistently across all the pooled samples and demonstrated significant expression differences between the AD and control groups.

### Expression of the validated AD biomarkers in patients’ blood

We analyzed the expression of the four AD RNA biomarkers in the single AD patients (n=50) and matching controls (n=50) utilizing RT-qPCR. All four AD biomarkers were significantly downregulated (*p* <0.05) in the peripheral blood of AD patients when compared to matching healthy controls (Fig. 3A). The difference between the average normalized Ct value in the control samples and the normalized Ct value in each AD sample for each of the 4 biomarkers was combined to obtain an AD predictive score (ADps) for each individual (Fig. 3B). To determine the relationship between the ADps and AD status, receiver operating characteristic (ROC) analysis was performed in the cohort of patients studied utilizing panels composed of either all four biomarkers or different combinations of three biomarkers. The area under the ROC curve (AUC) was similar for most biomarker combinations utilized in the analysis, except for the panel composed of TNRC18, PML and ZYX, which displayed a lower AUC when compared with the other biomarker combinations (AUC = 0.715 vs AUC >0.797) (Fig. 3B). Sensitivity and specificity were analyzed for the different combinations of the 4 biomarkers utilizing an ADps threshold of >1.5 or >1.25. The four-biomarker panel and the combination ADAMTSL4, PML, ZYX yielded the highest specificity and sensitivity (Fig. 3C), with an extremely high specificity (96%), a positive predictive value (PPV) of 92% and a negative predictive value (NPV) of 67% when an ADps threshold of >1.5 was applied (Fig. 3D).

### Relationship between ADps and other parameters

To examine whether the diagnostic value of the four RNA biomarker panel (ADps score) was influenced by demographic or clinical variables, including age, race, sex or MMSE score, we conducted correlation and group analyses using Spearman correlation, the Mann-Whitney U test, or Welch’s t-test. No significant correlation was found between the ADps and sex, race or MMSE score in either the AD or control groups (*p* >0.05, ρ <±0.1) (Fig. 4A,C,D,E). However, a significant inverse correlation was observed between ADps and age in the AD patients (*p* 0.0017, π -.44) but not in the control group (Fig. 4B). Altogether, these findings suggest that the four-biomarker, ADps score may have potential utility in detecting early-stage Alzheimer disease.

**Figure 4.**
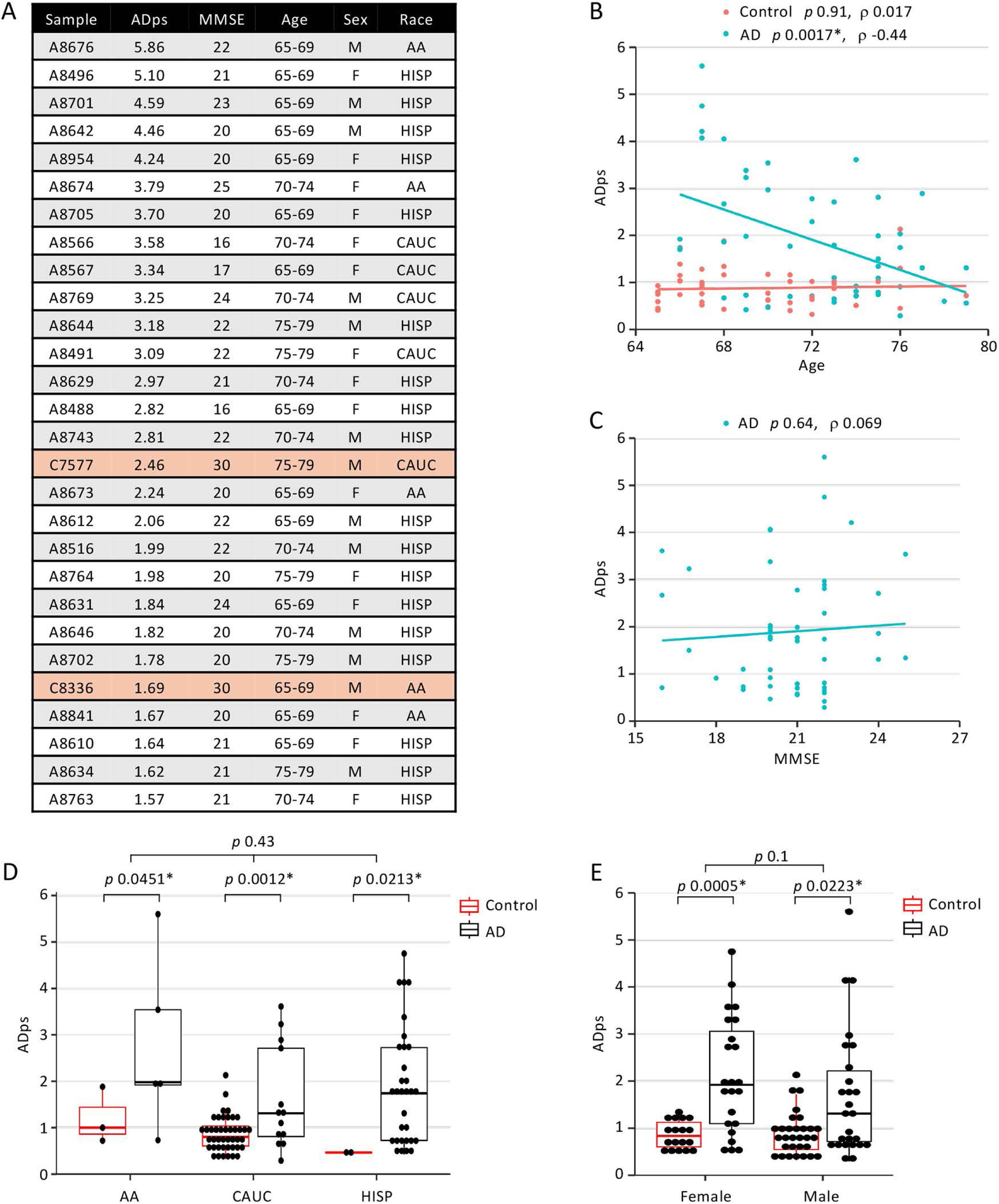
Correlation between the ADps from the 4 RNA biomarker panel and key patients’ clinical data. (A) Clinical data of AD patients and control subjects with an ADps >1.5. (B) Spearman correlation analyses between age and ADps in AD patients and control subjects. * *p* <0.05 was considered statistically significant. (C) Spearman correlation analysis between ADps and MMSE scores in AD patients and control subjects. (D) Correlation between race and ADps score assessed via 2-way ANOVA; *p*-value <0.05 was considered statistically significant. (E) Association between sex and ADps assessed via Mann-Whitney U test; *p* value <0.05 was considered statistically significant.

## Discussion

A whole transcriptome RNAseq approach was utilized to analyze RNAs expression levels and alternative splicing events in peripheral whole blood from AD patients and control subjects. From this analysis, 35 RNAs were selected for validation, leading to the identification of four coding mRNAs that formed a novel biomarker panel for a minimally invasive AD diagnosis. A novel AD predictive score (ADps) algorithm was developed to quantify the relative expression of the four biomarkers in individual AD patients compared to their average expression in the control population. Using a well-characterized cohort of 50 AD patients defined by clinical diagnosis and cognitive test scores, we showed that setting an ADps threshold of >1.5 yielded a positive predictive value (PPV) of the four-biomarker panel greater than 90% and a specificity exceeding 95%. These performance characteristics surpass those for most of the biomarkers currently studied and being developed as diagnostic tools [35–39].

The four RNA biomarkers identified in this study are encoded by genes with well-established functions in biological processes related to AD or other cognitive impairment disorders. ADAMTSL4 encodes an extracellular matrix glycoprotein that promotes fibrillin-1 microfibril assembly. Elevated expression of ADAMTSL4 has been associated with greater white matter hyperintensity burden in aging populations, suggesting its possible involvement in cerebrovascular changes linked to dementia [40]. PML encodes the promyelocytic leukemia protein, a key component of nuclear bodies that regulate transcription, protein turnover, and stress responses. PML is expressed in neurons and may mediate responses to proteins aggregates, such as the amyloid precursor protein (APP) fragment, which forms PML-containing nuclear aggregates that are reduced in AD hippocampi with high amyloid plaque load [41]. TNRC18 is a chromatin-associated protein that binds H3K9me3 and recruits transcriptional repressors to silence retrotransposons [42]. Although there’s no known direct link between TNRC18 and AD or other forms of dementia the potential role of retrotransposons reactivation in neural disease suggests that TNRC18 may play an indirect role in dementia and other neurological disorders. ZYX encodes zyxin, which links actin dynamics to intracellular signaling and gene regulation. Zyxin is downregulated in neuronal cells exposed to amyloid-beta [43], and proteomic analysis of neuron-derived exosomes isolated from blood samples identified it as a potential AD biomarker, with decreased protein levels observed in individuals with mild cognitive impairment (MCI) and AD [44]. Although the diagnostic performance of this assay remains to be fully validated, it is encouraging that the decline in the zyxin protein observed in the exosomes of a small number of AD patients (n=5) is consistent with the reduction in ZYX RNA levels we observed in blood samples from a larger number of AD patients (n=50).

One possible reason for the lack of effectiveness for the few existing therapies aimed at slowing the progression of AD and related disorders is the advanced stage of the disease at the time of diagnosis and pharmacological intervention. Thus, there is a need for biomarkers that can identify patients in earlier stages of the disease, with the goal of introducing therapies before the neurological damage becomes evident and is already irreversible. Although no correlation was observed between ADps score and race, sex or MMSE score in either AD or control subjects, a significant inverse correlation with age was observed in the AD patients but not in the control subjects. These data indicate that the four-biomarker panel may serve as a rapid, reliable, non-invasive tool for the early diagnosis of AD, particularly at stages when cognitive impairment is subtle and clinical evaluation alone may be inconclusive.

Many AD diagnostic approaches are limited by the need for specialized equipment, extensive operator training, and high costs, and often rely on biomarkers typically only detectable in CSF. In contrast, our integrated approach validates a panel of four RNAs that can be easily and reliably quantified using standard clinical lab equipment with minimal operator training. This method offers a minimally invasive, cost effective and scalable alternative for AD diagnosis. While this study analyzed a medium-sized cohort of patients and controls, it is plausible that individual variations in the expression of the RNAs we characterized might be more pronounced in the subjects analyzed, thus reducing the predictive power of the biomarker panel. Furthermore, since much of the clinical diagnosis is based on cognitive tests, it is possible that some patients included in the study may have cognitive impairment due to conditions other than AD. Going forward, a larger longitudinal study will be needed to further validate the four-biomarker panel as an early-stage AD diagnostic tool, assess its performance across diverse populations, and potentially refine the panel by reducing it to three biomarkers, without compromising its sensitivity and specificity, as suggested by our initial data.

## Conclusions

The data presented in this study indicate that a novel peripheral blood RNA biomarker panel can be used to diagnose Alzheimer’s disease with a specificity of >95% and a positive predictive value of >90%. Transcripts encoded by the genes ADAMTSL4, PML, TNRC18, and ZYX are differentially expressed in the peripheral blood of patients with Alzheimer’s disease and can be easily quantified for use as a minimally invasive, cost-effective, and reliable diagnostic tool that requires standard clinical laboratory equipment and minimal operator training. This four-RNA biomarker panel also has the potential to serve as an early diagnostic tool for Alzheimer’s disease.

## Data Availability

The datasets generated and analyzed in this study are available in the NIH Gene Expression Omnibus (GEO) repository. GSE248423, https://www.ncbi.nlm.nih.gov/geo/query/acc.cgi?acc=GSE248423.

## Authors’ contributions

MC organized and planned most of the study, analyzed the data and wrote most of the manuscript. SP carried out the RNA extractions and validation assays, analyzed the RNA sequencing data, carried out statistical analysis and contributed generating the figures and writing the manuscript. JDR helped in planning the study and the experimental approach and was a contributor in writing the manuscript. All authors read and approved the final manuscript.

## Ethics approval and consent to participate

Blood samples were obtained from a commercial biobank (PrecisionMed). Samples were de-identified and are exempt from Institutional Review Board (IRB) approval. Patient consent records and other identifiable personal data are kept by the biobank Precisionmed and cannot be accessed by the authors of this study.

## Consent for publication

Not applicable.

## Lead contact

Further information and requests for resources and reagents should be directed to and will be fulfilled by the lead contact, Massimo Caputi.

## Data Availability

All data produced in the present study are available upon reasonable request to the authors

