## Supplemental Figures and Tables for "An RNA biomarker panel for the diagnosis of Alzheimer’s disease from whole blood"

Figure S1.

Boxplot of the normalized Ct values obtained by amplifying the total RNA extracted from 50 AD patients and 50 healthy control subjects for two miRNAs not validated by the analysis of pooled RNAs. * *p* value (Mann-Whitney U test) >0.05 was considered significant.

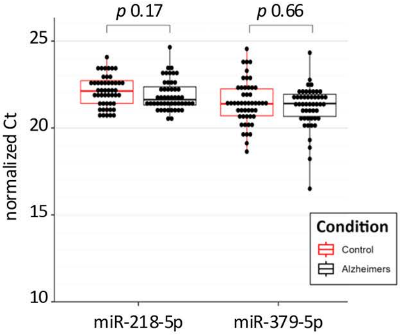

Figure S2.

Flowchart for the in-house and Novogene processing pipelines utilized for the analysis of the transcriptome sequencing raw reads for the identification of DE: i) mRNAs, ii) lncRNAs, iii) miRNAs, v) circRNAs and vi) alternatively spliced mRNA isoforms. Tools utilized in the In-house and Novogene pipelines are shown.

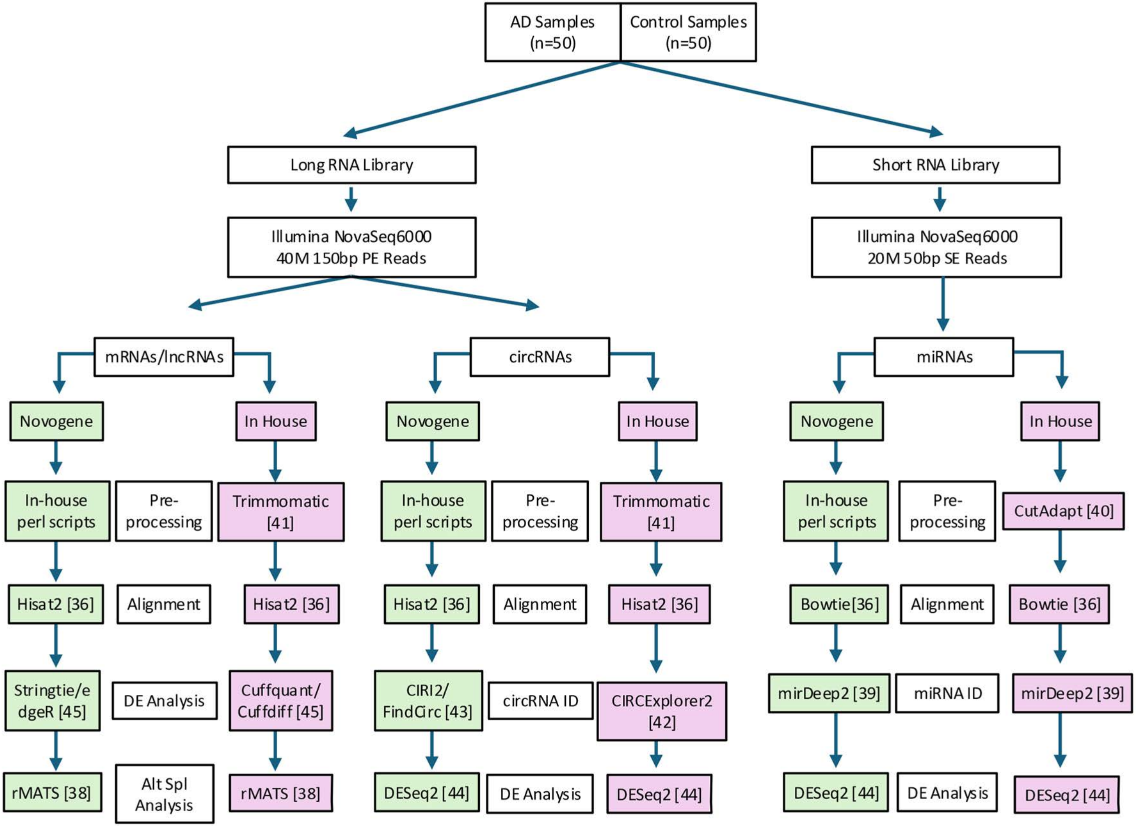

Table S1
AD patients and healthy control subjects' clinical data.

| Sample | Sample Type | MMSE | ADAS-Cog | Age | Sex | Race | Pooled RNA group |
|---|---|---|---|---|---|---|---|
| A8355 | AD | 17 | 44 | 75-79 | M | CAUC | Pooled 1AD |
| A8362 | AD | 24 | 28 | 75-79 | M | CAUC | Pooled 1AD |
| A8412 | AD | 16 | 38 | 70-74 | F | CAUC | Pooled 1AD |
| A8420 | AD | 19 | 27 | 70-74 | F | CAUC | Pooled 1AD |
| A8464 | AD | 18 | 31 | 75-79 | F | CAUC | Pooled 1AD |
| A8488 | AD | 16 | 46 | 65-69 | F | HISP | Pooled 2AD |
| A8491 | AD | 22 | 21 | 75-79 | F | CAUC | Pooled 2AD |
| A8496 | AD | 22 | 27 | 65-69 | F | HISP | Pooled 2AD |
| A8500 | AD | 22 | 27 | 70-74 | M | CAUC | Pooled 2AD |
| A8515 | AD | 20 | 30 | 75-79 | M | HISP | Pooled 2AD |
| A8516 | AD | 22 | 33 | 70-74 | M | HISP | Pooled 3AD |
| A8547 | AD | 22 | 46 | 75-79 | M | HISP | Pooled 3AD |
| A8566 | AD | 16 | 43 | 70-74 | F | CAUC | Pooled 3AD |
| A8567 | AD | 17 | 33 | 65-69 | F | CAUC | Pooled 3AD |
| A8603 | AD | 21 | 32 | 70-74 | F | HISP | Pooled 3AD |
| A8610 | AD | 21 | 38 | 65-69 | F | HISP | Pooled 4AD |
| A8612 | AD | 22 | 41 | 65-69 | M | HISP | Pooled 4AD |
| A8629 | AD | 21 | 39 | 70-74 | F | HISP | Pooled 4AD |
| A8631 | AD | 24 | 36 | 65-69 | F | HISP | Pooled 4AD |
| A8634 | AD | 21 | 37 | 75-79 | M | HISP | Pooled 4AD |
| A8642 | AD | 20 | 38 | 65-69 | M | HISP | Pooled 5AD |
| A8644 | AD | 22 | 39 | 75-79 | M | HISP | Pooled 5AD |
| A8646 | AD | 20 | 32 | 70-74 | M | HISP | Pooled 5AD |
| A8655 | AD | 25 | 21 | 75-79 | F | CAUC | Pooled 5AD |
| A8669 | AD | 19 | 25 | 65-69 | M | AA | Pooled 5AD |
| A8673 | AD | 20 | 24 | 65-69 | F | AA | Pooled 6AD |
| A8674 | AD | 25 | 17 | 70-74 | F | AA | Pooled 6AD |
| A8676 | AD | 22 | 17 | 65-69 | M | AA | Pooled 6AD |
| A8701 | AD | 23 | 32 | 65-69 | M | HISP | Pooled 6AD |
| A8702 | AD | 20 | 32 | 75-79 | M | HISP | Pooled 6AD |
| A8705 | AD | 20 | 31 | 65-69 | F | HISP | Pooled 7AD |
| A8716 | AD | 22 | 16 | 75-79 | M | CAUC | Pooled 7AD |
| A8738 | AD | 21 | 31 | 75-79 | M | HISP | Pooled 7AD |
| A8739 | AD | 20 | 33 | 70-74 | M | HISP | Pooled 7AD |
| A8740 | AD | 19 | 36 | 65-69 | M | HISP | Pooled 7AD |
| A8741 | AD | 22 | 33 | 65-69 | M | HISP | Pooled 8AD |
| A8742 | AD | 22 | 34 | 70-74 | M | HISP | Pooled 8AD |
| A8743 | AD | 22 | 36 | 70-74 | M | HISP | Pooled 8AD |

| | | | | | | | |
|---|---|---|---|---|---|---|---|
| A8750 | AD | 22 | 15 | 65-69 | F | AA | Pooled 8AD |
| A8758 | AD | 20 | 39 | 70-74 | F | HISP | Pooled 8AD |
| A8759 | AD | 21 | 35 | 75-79 | M | HISP | |
| A8761 | AD | 21 | 40 | 70-74 | M | HISP | |
| A8762 | AD | 22 | 39 | 70-74 | M | HISP | |
| A8763 | AD | 21 | 43 | 70-74 | F | HISP | |
| A8764 | AD | 20 | 41 | 75-79 | F | HISP | |
| A8766 | AD | 20 | 41 | 75-79 | F | HISP | |
| A8768 | AD | 22 | 26 | 75-79 | F | CAUC | |
| A8769 | AD | 24 | 21 | 70-74 | M | CAUC | |
| A8841 | AD | 20 | 24 | 65-69 | F | AA | |
| A8954 | AD | 20 | 31 | 65-69 | F | HISP | |
| C7090084 | Control | 30 | | 70-74 | M | CAUC | Pooled 1C |
| C7090092 | Control | 30 | | 70-74 | F | CAUC | Pooled 1C |
| C7508 | Control | 29 | | 70-74 | M | HISP | Pooled 1C |
| C7529 | Control | 30 | | 65-69 | M | CAUC | Pooled 1C |
| C7534 | Control | 29 | | 70-74 | F | CAUC | Pooled 1C |
| C7535 | Control | 29 | | 65-69 | F | CAUC | Pooled 2C |
| C7541 | Control | 29 | | 70-74 | M | CAUC | Pooled 2C |
| C7564 | Control | 30 | | 65-69 | M | CAUC | Pooled 2C |
| C7568 | Control | 29 | | 70-74 | F | CAUC | Pooled 2C |
| C7571 | Control | 30 | | 70-74 | M | CAUC | Pooled 2C |
| C7577 | Control | 30 | | 75-79 | M | CAUC | Pooled 3C |
| C7578 | Control | 29 | | 70-74 | M | CAUC | Pooled 3C |
| C7580 | Control | 30 | | 65-69 | F | CAUC | Pooled 3C |
| C7581 | Control | 30 | | 65-69 | F | CAUC | Pooled 3C |
| C7585 | Control | 30 | | 65-69 | M | CAUC | Pooled 3C |
| C7591 | Control | 29 | | 75-79 | F | AA | Pooled 4C |
| C7592 | Control | 29 | | 65-69 | F | CAUC | Pooled 4C |
| C7594 | Control | 29 | | 75-79 | M | CAUC | Pooled 4C |
| C7595 | Control | 28 | | 65-69 | M | HISP | Pooled 4C |
| C7596 | Control | 29 | | 70-74 | F | CAUC | Pooled 4C |
| C7597 | Control | 30 | | 65-69 | M | CAUC | Pooled 5C |
| C7598 | Control | 30 | | 70-74 | M | CAUC | Pooled 5C |
| C7601 | Control | 29 | | 65-69 | M | AA | Pooled 5C |
| C7602 | Control | 28 | | 75-79 | F | CAUC | Pooled 5C |
| C7603 | Control | 30 | | 70-74 | F | CAUC | Pooled 5C |
| C7604 | Control | 30 | | 65-69 | M | CAUC | Pooled 6C |
| C7605 | Control | 30 | | 65-69 | M | CAUC | Pooled 6C |
| C7606 | Control | 29 | | 65-69 | M | CAUC | Pooled 6C |
| C7607 | Control | 30 | | 70-74 | M | CAUC | Pooled 6C |
| C8201 | Control | 30 | | 75-79 | F | CAUC | Pooled 6C |
| C8202 | Control | 30 | | 70-74 | M | CAUC | Pooled 7C |

| | | | | | | | |
|---|---|---|---|---|---|---|---|
| C8204 | Control | 30 | | 70-74 | M | CAUC | Pooled 7C |
| C8206 | Control | 30 | | 65-69 | M | CAUC | Pooled 7C |
| C8207 | Control | 30 | | 65-69 | F | CAUC | Pooled 7C |
| C8208 | Control | 29 | | 65-69 | M | CAUC | Pooled 7C |
| C8210 | Control | 30 | | 65-69 | M | CAUC | Pooled 8C |
| C8211 | Control | 30 | | 65-69 | F | CAUC | Pooled 8C |
| C8212 | Control | 30 | | 70-74 | F | CAUC | Pooled 8C |
| C8213 | Control | 30 | | 70-74 | M | CAUC | Pooled 8C |
| C8214 | Control | 29 | | 70-74 | M | CAUC | Pooled 8C |
| C8216 | Control | 30 | | 65-69 | M | CAUC | |
| C8217 | Control | 29 | | 65-69 | M | CAUC | |
| C8218 | Control | 30 | | 65-69 | F | CAUC | |
| C8222 | Control | 30 | | 65-69 | F | CAUC | |
| C8231 | Control | 30 | | 70-74 | F | CAUC | |
| C8237 | Control | 30 | | 65-69 | M | CAUC | |
| C8252 | Control | 29 | | 70-74 | M | CAUC | |
| C8260 | Control | 30 | | 65-69 | M | CAUC | |
| C8324 | Control | 30 | | 65-69 | M | CAUC | |
| C8336 | Control | 30 | | 65-69 | M | AA | |

Table S2
Sequence of the primers utilized for qPCR, RT-PCR. The primer target and type of assay are indicated.

| **Primer Name** | **Sequence (5'-3')** | **Target** | **Assay** |
|---|---|---|---|
| qPCR-RPL13A-F | AGCCAGAAGACTGATTGGAG | RPL13A | RT-qPCR |
| qPCR-RPL13A-R | AGTGCTTGCACATTCTAACAG | RPL13A | RT-qPCR |
| qPCR-ADAMTSL4-F | AGAGGAGACCCAGGAGATTC | ADAMTSL4 | RT-qPCR |
| qPCR-ADAMTSL4-R | AAAGGGCACTCTCCCATAAC | ADAMTSL | RT-qPCR |
| qPCR-BEAN1-F | CTGTTCTGCTCCAAGGATTAC | BEAN1 | RT-qPCR |
| qPCR-BEAN1-R | TCCGAGCTCTCTGAGAATGTG | BEAN1 | RT-qPCR |
| qPCR-DNAH17-F | CCTCGCTGCTCAGTACATAAA | DNAH17 | RT-qPCR |
| qPCR-DNAH17-R | TCATTGATCAGCACCAGAAAC | DNAH17 | RT-qPCR |
| qPCR-PML-F | AGTCAGGAACTAGAGACCAG | PML | RT-qPCR |
| qPCR-PML-R | GTCACTCAAGGAACACAGAG | PML | RT-qPCR |
| qPCR-TNRC18-F | GACTTGATCACCGTGGAGTTTG | TNRC18 | RT-qPCR |
| qPCR-TNRC18-R | CTCAGCACACTGTATCTTATAG | TNRC18 | RT-qPCR |
| qPCR-PEAK3-F | TCCCAAAGTGCTGGGATTAC | PEAK3 | RT-qPCR |
| qPCR-PEAK3-R | CAGAGTGGGCACCTTTATTTG | PEAK3 | RT-qPCR |
| qPCR-ZYX-F | AGCCTGTGTCTTTGGCTAAC | ZYX | RT-qPCR |
| qPCR-ZYX-R | TGGAAGCCACAGGAGTAAAC | ZYX | RT-qPCR |
| qPCR-miR-9-5p | CTTTGGTTATCTAGCTGTATG | miR-9-5p | RT-qPCR |
| qPCR-miR-708-3p | ACTAGACTGTGAGCTTCTAG | miR-708-3p | RT-qPCR |
| qPCR-miR-411-5p | TAGTAGACCGTATAGCGTAC | miR-411-5p | RT-qPCR |
| qPCR-miR-379-5p | TGGTAGACTATGGAACGTAG | miR-379-5p | RT-qPCR |
| qPCR-miR-302-p | TAAGTGCTTCCATGTTTGAG | miR-302-p | RT-qPCR |
| qPCR-miR-218-5p | TTGTGCTTGATCTAACCATG | miR-218-5p | RT-qPCR |
| qPCR-miR-203a-3p | GAAATGTTTAGGACCACTAG | miR-203a-3p | RT-qPCR |
| qPCR-miR-200b-3p | AATACTGCCTGGTAATGATG | miR-200b-3p | RT-qPCR |
| qPCR-miR-129-5p | CTTTTTGCGGTCTGGGCTTG | miR-129-5p | RT-qPCR |
| qPCR-miR-532 | TCTGACTCTCCGCCTCTCC | miR-532 | RT-qPCR |
| 5'U6a | GCTTCGGCAGCACATATACTA | U6 snRNA | RT-qPCR |
| 3'U6a | CGAATTTGCGTGTCATCCTT | U6 snRNA | RT-qPCR |
| qPCR-FAM135A-F | TGACATGTCGAGATCACTCAG | FAM135A_0007 | RT-qPCR |
| qPCR-FAM135A-R | CAATCCAAGTTCAATGTAAG | FAM135A_0007 | RT-qPCR |
| qPCR-SPATA13-F | CAGTACAACAAAGAGGAACCTC | SPATA13_0008 | RT-qPCR |
| qPCR-SPATA13-R | CTAGCTGCGCAACGGTGAACA | SPATA13_0008 | RT-qPCR |
| RPL17-F | ACGCAGAGAGTAATGCTGAAC | RPL17 | RT-PCR |
| RPL17-R | AATTTACTCCCGTGCCATAAG | RPL17 | RT-PCR |
| GIGYF2-F | GGTTCTGAGACTCCCTGTC | GIGYF2 | RT-PCR |
| GIGYF2-R | GTTGTCTTTAAGGAAAAGTG | GIGYF2 | RT-PCR |

| | | | |
|---|---|---|---|
| WWP2-F | AGGAGTTCGGCGCCGCTTCTG | WWP2 | RT-PCR |
| WWP2-R | ATGATCTCATTCCAGAGAAG | WWP2 | RT-PCR |
| FXYD5-F | GACGTTGAAAGATACCACGTC | FXYD5 | RT-PCR |
| FXYD5-R | ATGATGATGCCTGTGATGAAC | FXYD5 | RT-PCR |
| ZDHHC4-F | AGTCTCGTATCGCGCCCGGGAG | ZDHHC4 | RT-PCR |
| ZDHHC4-R | CAAGCTATGGGTTTTCGAGCAG | ZDHHC4 | RT-PCR |
| SNRNP70-F | TACACATGGTCTACAGTAAG | SNRNP70 | RT-PCR |
| SNRNP70-R | GTGTCATCGCGGCCTGAATG | SNRNP70 | RT-PCR |
| TECPR1-F | AGGACCTACCACCGCGACTTC | TECPR1 | RT-PCR |
| TECPR1-R | TCCTGTACCGGATCCACTTC | TECPR1 | RT-PCR |
| TMEM104-F | CTTGGGAGCTGGCTGTGCTG | TMEM104 | RT-PCR |
| TMEM104-R | CACCCGGTCTGTGATTTCAAAC | TMEM104 | RT-PCR |
| RBIS-F | CACGCAAGTGTTAAACTTCTG | RBIS | RT-PCR |
| RBIS-R | CTTTCTGCAGAGGTTCAAGTG | RBIS | RT-PCR |
| PIGO-F | TGATTCCTCAAGTTAGCCTTG | PIGO | RT-PCR |
| PIGO-R | TGGCTCCCACTAGCAAAGCAG | PIGO | RT-PCR |
| MBD6-F | CAATGTTTGCAGAGATGTTCAG | MBD6 | RT-PCR |
| MBD6-R | GACATTAAGTGGACACTCCAG | MBD6 | RT-PCR |
| KDM2B-F | ACGGAGATGAGCATGTCCCAG | KDM2B | RT-PCR |
| KDM2B-R | ATAAGCATCGACTCCCTCTG | KDM2B | RT-PCR |
| CFAP44-F | GAGATCCACTGGAAAGTGATAC | CFAP44 | RT-PCR |
| CFAP44-R | AGAATCTAGGTGTCCTAGTTC | CFAP44 | RT-PCR |
| PICALM-F | GAAGTGTTCCTGTAATGACG | PICALM | RT-PCR |
| PICALM-R | CTTTATAAACTGTATTGATAC | PICALM | RT-PCR |
| PILRB-F | GCTGGCTAGGACTCCAGTAC | PILRB | RT-PCR |
| PILRB-R | GTGTCCAGCTCGACTCGGCAG | PILRB | RT-PCR |
